# A 360º View for Large Language Models: Early Detection of Amblyopia in Children using Multi-View Eye Movement Recordings

**DOI:** 10.1101/2024.05.03.24306688

**Authors:** Dipak P. Upadhyaya, Aasef G. Shaikh, Gokce Busra Cakir, Katrina Prantzalos, Pedram Golnari, Fatema F. Ghasia, Satya S. Sahoo

## Abstract

Amblyopia is a neurodevelopmental visual disorder that affects approximately 3-5% of children globally and it can lead to vision loss if it is not diagnosed and treated early. Traditional diagnostic methods, which rely on subjective assessments and expert interpretation of eye movement recordings presents challenges in resource-limited eye care centers. This study introduces a new approach that integrates the Gemini large language model (LLM) with eye-tracking data to develop a classification tool for diagnosis of patients with amblyopia. The study demonstrates: (1) LLMs can be successfully applied to the analysis of fixation eye movement data to diagnose patients with amblyopia; and (2) Input of medical subject matter expertise, introduced in this study in the form of medical expert augmented generation (MEAG), is an effective adaption of the generic retrieval augmented generation (RAG) approach for medical applications using LLMs. This study introduces a new multi-view prompting framework for ophthalmology applications that incorporates fine granularity feedback from pediatric ophthalmologist together with in-context learning to report an accuracy of 80% in diagnosing patients with amblyopia. In addition to the binary classification task, the classification tool is generalizable to specific subpopulations of amblyopic patients based on severity of amblyopia, type of amblyopia, and with or without nystagmus. The model reports an accuracy of: (1) 83% in classifying patients with moderate or severe amblyopia, (2) 81% in classifying patients with mild or treated amblyopia; and (3) 85% accuracy in classifying patients with nystagmus. To the best of our knowledge, this is the first study that defines a multiview prompting framework with MEAG to analyze eye tracking data for the diagnosis of amblyopic patients.

## 1. Introduction

Amblyopia is a neurodevelopmental disorder of the eye affecting 3-5% of the children’s population, that is, an estimated 99.2 million children are affected by amblyopia worldwide [1]. This condition leads to decreased vision in one eye due to improper visual stimulation during early childhood [1]. If left untreated, amblyopia can result in irreversible visual impairment, and it contributes to lifelong consequences for the affected individuals. In the United States, amblyopia is the most common cause of monocular vision loss among children, adolescents, and middle-aged adults, underscoring the substantial public health challenge it poses across age groups [1]. The diagnosis of amblyopia has traditionally relied on subjective visual acuity assessments and the interpretation of eye movement recordings, which requires specialized equipment and expert clinical analysis. These diagnostic challenges are exacerbated in resource-limited settings, where access to specialized pediatric ophthalmology services is often unavailable. The increasing use of high-fidelity eye tracking instruments, such as video oculography tracker (EyeLink1000 plus), has enabled the use of eye tracking data for the detection of amblyopia. In this study, we leverage the powerful transfer learning capabilities of the Gemini large language model (LLM) (Google, Inc.) to analyze eye tracking data in a cohort of 135 subjects to develop a high accuracy classification tool that can be deployed in remote, resource scarce eye care clinics.

LLMs have demonstrated state of the art performance in a variety of applications [2-5]; however, their performance in medical applications have highlighted multiple challenges due to the complexity of the medical domain [6-9]. Specifically, customized LLMs such as Med-PaLM, which extends the Pathway Language Model (PaLM) [3] and MedGemini [10], require the development of fine-tuned prompting methods to answer medical domain questions (e.g., questions in the MultiMedQA multiple-choice benchmark dataset). Building on the PaLM model, Gemini is a state-of-the-art language model that has achieved high accuracy in a range of natural language processing tasks [2, 11]. However, initial results from our approach to use the Gemini model to classify patients with amblyopia using eye movement data visualized as a cartesian plot resulted in low accuracy with multiple errors. Specifically, as a baseline prompt, we used eye tracking plots (Figure 1) corresponding to a single eye together with a textual description of the plot for the Gemini model. The model reported an accuracy of 50% or less for classifying patients with amblyopia with errors in classifying patients based on the severity of amblyopia and amblyopic patients with or without nystagmus.

**Figure 1:**
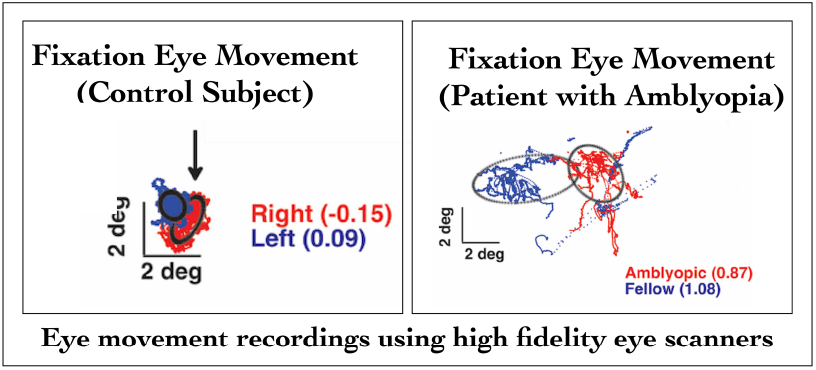
The visualization of eye movement recordings in children with amblyopia and control subjects with dense, focused saccades seen in control subjects (left); and dispersed, wide-ranging saccades seen in amblyopic patient (right)

To address these limitations, we built on the few shot learning capabilities of LLMs together with the development of a new multi-view prompting framework for ophthalmology applications. Specifically, we combine eye tracking data recorded under multiple viewing conditions to create a collage of eye movement recordings for each patient. In the next step, we systematically label the images in the collage with input from a pediatric ophthalmologist (co-author FG) and construct a structured prompt expression for each patient. Together with few-shot learning approach, this new multiple view prompt with medical expert feedback demonstrates:

1. The feasibility of using LLMs to classify patients with amblyopia using recordings from eye tracking instruments
2. The increase in contextual information (represented as a multi-view collage of eye movement plots in this study) is effective in improving the accuracy of results generated by language models in a specialized medical domain.

In contrast to fine tuning LLMs, which requires retraining the LLM to modify gradient weights, prompt tuning is a low-cost approach to improve the accuracy of LLM generated results [6]. However, prompt tuning for medical applications requires customizations that reflect the specific attributes of the medical data. Therefore, medical expertise augmented retrieval (MEAG), which is a form of retrieval augmented generation (RAG) [12] is essential for effective use of language models to analyze patient data. We discuss the characteristics of eye tracking data and its use in diagnosis of patients with amblyopia.

### 1.1 Background

Eye movement abnormalities are being increasingly used as diagnostic markers in amblyopia research as they can be recorded in a non-invasive manner [13]. Individuals with amblyopia demonstrate notable fixation instability, particularly in the amblyopic eye as compared to its fellow eye (Figure 1), which highlights the close link between fixation eye movement (FEM) abnormalities and the sensory deficits in amblyopia [14, 15]. The use of modern eye trackers provides an objective dataset crucial for assessing visual function and understanding the nuance of impairments in eye movement control associated with the disorder [16]. In parallel, the development of LLMs with capabilities to analyze multimodal data including text and images, presents new avenues for ophthalmology applications. Therefore, this study uses the Gemini LLM to analyze eye tracking data for classification of patients with amblyopia.

### 1.2. Related Work

The application of machine learning algorithms in ophthalmology, particularly for amblyopia detection, has evolved over time [17]. Convolutional neural networks (CNNs) have been employed to analyze retinal images for amblyogenic risk factors and they have shown greater accuracy as compared to traditional methods [18]. Other machine learning methods have been explored for classification of visual field defects and assessing binocular vision, which is important for amblyopia diagnosis [19]. However, traditional approaches require extensive training data and their effectiveness have been affected due to the difficulty in identifying subtle features associated with amblyopia [20]. We have addressed some of these challenges in this study by leveraging the powerful capabilities of LLMs, including transfer learning that rely on few-shot based incontext learning for generating accurate results. In-context learning refers to the ability of language models to adapt to new tasks or domains without explicit retraining of the LLM through the use of one or more examples [2, 21].

## 2. Method

Figure 2 gives an overview of the method used in this study that consists of a multistage workflow, including: (i) acquisition of eye movement data using high fidelity eye tracking instrument; (ii) signal pre-processing methods to enhance data quality by reducing noise in the recordings; (iii) visualization of eye tracking recordings with appropriate scaling measures as cartesian plots; and (iv) the use of multiview prompting with input from pediatric ophthalmologist.

**Figure 2:**
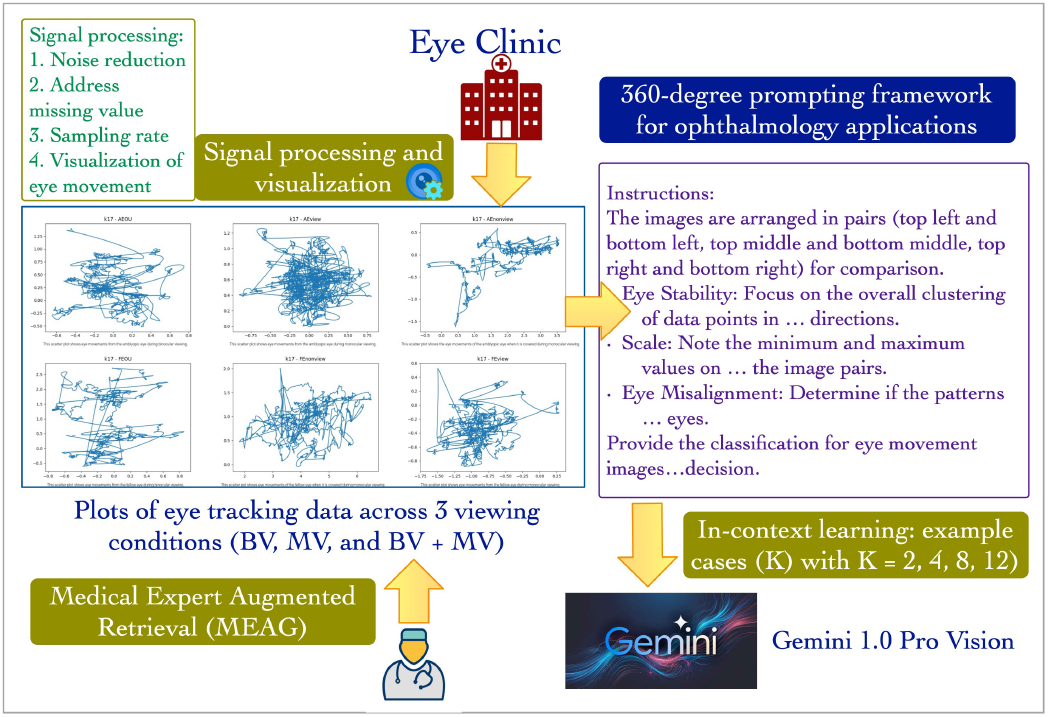
An overview of this study that introduced a new Multiview imaging LLM prompting techniques for analyzing pediatric ophthalmology data.

### 2.1 Study Cohort and Data Collection

This study uses eye movement recordings from 135 participants collected at the Cleveland Clinic Eye Institute (the study was reviewed and approved by the Cleveland Clinic Institutional Review Board). The study cohort consisted of 95 patients with amblyopia and 40 control subjects. The age of the participants ranged between 3 years and 72 years. Control subjects were selected based on the absence of ocular or systemic abnormalities affecting visual acuity, except for refractive errors. The 95 patients with amblyopia included 51 females and 44 males. The 40 control subjects included 23 females and 17 males. The control group consisted of 24 White Caucasian, 6 African American, 1 Hispanic, 5 Asians, and 4 multi-cultural patients. The amblyopic subjects included 61 Whites Caucasians, 12 African American, 10 Hispanics, 3 Asians, and 9 multi-cultural patients.

The EyeLink1000 plus video-based eye tracker was used with a sampling rate of 500 Hz with participants wearing corrective lenses. In a dimly lit room, participants faced an LCD screen displaying various stimuli, with calibration involving a cartoon animal accompanied by sound cues to focus attention. The calibration process adhered to a 5-point manufacturer guideline [22]. The fixation measurements were taken as subjects focused on a white dot against a black screen, employing animal sounds to redirect attention if needed. Each session was recorded with both eyes open (labeled as binocular vision, BV) or with left/right eye viewing while the other was covered (labeled as monocular viewing, MV), in a sequence determined randomly. The MV sessions were recorded from the active viewing eye, distinguishing between fellow eye (FE) and amblyopic eye viewing (AE) conditions. The covered eye was monitored using an infrared filter to avoid exposure to visible light.

### 2.2 A Multi-view Prompting Framework with Few-Shot Learning

Following the data pre-processing steps, the eye tracking data were visualized as cartesian plots using the Pandas library for data processing and the Python Imaging Library (PIL). As described in the previous section, the plots of eye tracking data were combined into a collage of six images with recording of each eye under three viewing conditions, that BV, MV, and BV + MV. In the next step, the collage was annotated with text prepared by pediatric ophthalmologist to describe the characteristics of the eye tracking data (Figure 3). In the final stage, the prompt expression was constructed using this multi-view expert annotated images together with structured input that described the context of the task and added example query examples as part of the few-shot learning approach.

**Figure 3:**
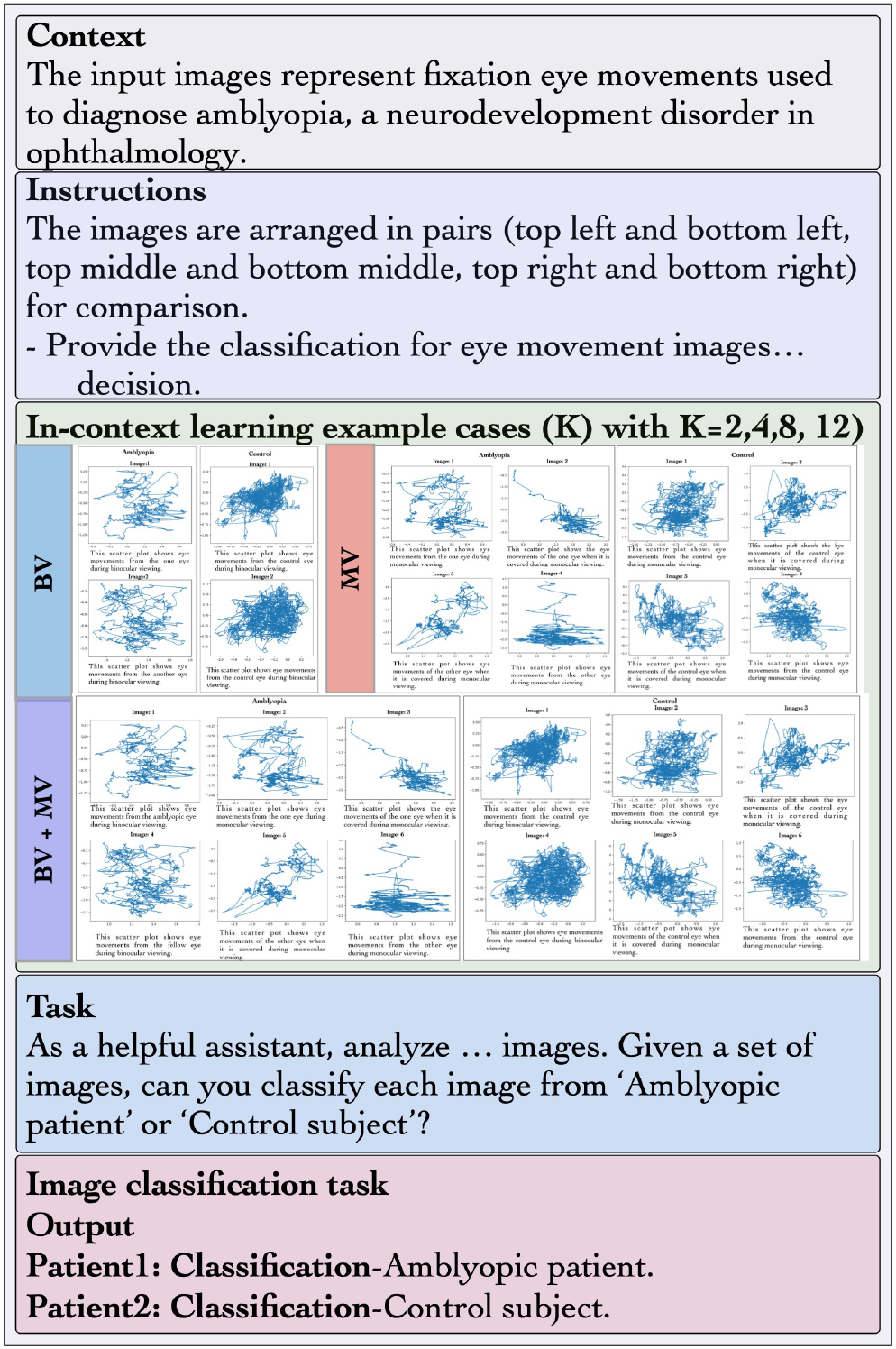
Overview of multi-view prompting framework developed for diagnosis of amblyopia using structured feedback from pediatric ophthalmologist.

### 2.3 In-context Learning for Eye Movement Data

LLMs have demonstrated a notable capability to learn specific contexts using few examples (few-shot or in-context learning) that was highlighted in the Generative Pretrained Transformer (GPT 3.5) model and many other subsequent studies [2, 3, 6]. In this study, we evaluated the performance of the Gemini LLM by combining the multiview prompt expressions with medical expertise together with in-context learning using a different number of examples for in-context learning. Figure 3 shows an overview of the multi-view prompting approach developed and implemented in this study for the Gemini LLM. This prompting framework includes several constituent tasks, including the system prompt together with multi-view expert annotated text and examples for few-shot in-context learning. Using this new prompting framework, the classification tool was evaluated using held out test dataset with 123 subjects (26 control subjects and 87 patients with amblyopia). We evaluated the effectiveness of the new multi-view prompting approach with and without feedback from the medical expert together with a systematic evaluation of the impact of different number of examples on the accuracy of the model.

## 3. Results

### 3.1 Experimental Setup

This study used Gemini 1.0 Pro Vision model that accepts both text and images as input and generates text as output [4]. The evaluation was structured into four few-shot prompting scenarios with multi-view collages corresponding to recordings from 2, 4, 8 or 12 subjects under the three viewing conditions. The subjects used in the few-shot prompting were selected to be representative of amblyopic patients and control subjects. This evaluation process was performed for each of the three viewing conditions (BV, MV, and BV + MV).

To demonstrate the effectiveness of prompt expression created using fine granularity input from pediatric ophthalmologist, we performed a comparative evaluation of the LLM with and without the inclusion of medical expert generated text. The results of this evaluation demonstrated the importance of feedback from medical experts in constructing effective prompts. Based on these results, all subsequent prompts incorporate the input of ophthalmologist in labeling the eye movement plots. In the second phase of the evaluation, the prompt-tuned Gemini model was used to classify patients with amblyopia based on the severity of amblyopia, that is, across the three categories of: (1) patients with severe or moderate amblyopia and control subjects; (2) patients with mild or treated amblyopia and control subjects; and (3) amblyopic patient with nystagmus and control subjects. A key feature of the multi-view prompt development was an iterative feedback mechanism that tuned the prompt to reflect the reasoning approach used by pediatric ophthalmologists in diagnosing patients with amblyopia.

The evaluation experiments were conducted on a server with 32 GB RAM and Intel® Core™ i7-13700K CPU, using Ubuntu 22.04.1 LTS (64-bit). The 123 subjects constituting the test dataset did not have any overlap with the 12 subjects used for incontext learning. Temperature is a tunable parameter of LLMs that corresponds to increased randomness in results with higher temperature value and more deterministic results with lower temperature values [7]. In this study, a temperature parameter of 0.7 was found to be optimal for achieving the highest accuracy.

### 3.2. Evaluation Results

Table 1 lists the performance of the LLM in phase 1 using a single image corresponding to eye movement recordings under a specific viewing condition. The results generated by the Gemini model have low accuracy due to the limited context provided in the prompt, including a limited “view” of the eye movement recording and lack of fine granularity medical expert feedback in the prompt. Specifically, the model reports an accuracy of 0.54 when distinguishing between patients with amblyopia (irrespective of their severity) and control subjects; an accuracy of 0.58 for diagnosing patients with mild or treated amblyopia; and an accuracy of 0.62 for diagnosing patients with moderate and severe amblyopia.

**Table 1.** Performance of the classification tool using a baseline prompt to: (1) classify patients with amblyopia and control subjects; (2) diagnosing patients based on their severity.

| Model | Amblyopia versus control (BV) | Mild and treated amblyopia versus control (BV) | Moderate and severe amblyopia versus control (BV) |
| --- | --- | --- | --- |
| **Few shot (K=12)** | 0.54 | 0.58 | 0.62 |

To address the limitations of the baseline prompting approach, we used the multi-view prompting approach as described in the previous section (Section 2.2). Table 2 shows the evaluation results that compares the performance of the Gemini model with and without feedback from the medical expert. Table 2 lists the results across the three different viewing conditions with few-shot learning approach. The results validate the effectiveness of few-shot learning in LLMs with improvement in performance across the three viewing conditions with increasing number of examples. Further, the results show that there is a significant improvement in the performance of the model with input of the medical expert.

**Table 2.** Comparative evaluation of the classification tool with and without MEAG prompting approach for classifying amblyopic patients and control subjects across the three viewing conditions.

| Model | BV + MV viewing condition (baseline prompt) Accuracy | BV + MV viewing condition (MEAG prompt) Accuracy | MV viewing condition (baseline prompt) Accuracy | MV viewing condition (MEAG prompt) Accuracy | BV viewing condition (baseline prompt) Accuracy | BV viewing condition (MEAG prompt) Accuracy |
| --- | --- | --- | --- | --- | --- | --- |
| **Few shot (K=2)** | 0.55 | 0.62 | 0.55 | 0.63 | 0.6 | 0.65 |
| **Few shot (K=4)** | 0.55 | 0.64 | 0.55 | 0.67 | 0.6 | 0.6 |
| **Few shot (K=8)** | 0.57 | 0.74 | 0.57 | 0.79 | 0.62 | 0.79 |
| **Few shot (K=12)** | 0.57 | 0.78 | 0.57 | 0.78 | 0.62 | 0.8 |

*BV: Binocular viewing (Two images per collage), MV: Monocular viewing (Four images per collage), BV+MV (6 images per collage)*

Table 3 lists the performance of the models across different classification tasks based on: (1) the severity of amblyopia, including mild and treated amblyopia, moderate and severe amblyopia, and (2) amblyopia with nystagmus as compared to the control group using the few-shot in-context learning approach. Table 3 also includes the results for combined binocular and monocular viewing (BV + MV), MV, and BV models, which demonstrates the improvement in accuracy of the model with an increased number of examples for in-context learning across all viewing conditions. We also note that the classification accuracy is the highest for eye tracking data recorded under the binocular viewing condition.

**Table 3.** An evaluation of the generalizability capabilities of the classification tool using multiview prompting framework with MEAG across different subpopulations of amblyopic patients.

| Model | BV + MV viewing condition Accuracy | MV viewing condition Accuracy | BV viewing condition Accuracy |
| --- | --- | --- | --- |
| **Mild and treated amblyopia versus control** |  |  |  |
| Few shot (K=2) | 0.6 | 0.65 | 0.7 |
| Few shot (K=4) | 0.63 | 0.69 | 0.68 |
| Few shot (K=8) | 0.76 | 0.78 | 0.8 |
| Few shot (K=12) | 0.77 | 0.81 | 0.81 |
| **Moderate and severe amblyopia versus control** |  |  |  |
| Few shot (K=2) | 0.68 | 0.69 | 0.7 |
| Few shot (K=4) | 0.71 | 0.74 | 0.77 |
| Few shot (K=8) | 0.78 | 0.8 | 0.81 |
| Few shot (K=12) | 0.8 | 0.82 | 0.83 |
| **Amblyopia with nystagmus versus control** |  |  |  |
| Few shot (K=2) | 0.7 | 0.72 | 0.73 |
| Few shot (K=4) | 0.74 | 0.74 | 0.76 |
| Few shot (K=8) | 0.8 | 0.81 | 0.83 |
| Few shot (K=12) | 0.8 | 0.83 | 0.85 |

## 4. Discussion and Conclusion

The study defines a new prompting framework for use with foundational language models in ophthalmology applications. The multi-view prompting framework that provides an enhanced visual context to language model is effective in leveraging the transfer learning capabilities of language model to accurately analyze eye tracking data. Further, the results demonstrate that notwithstanding the few shot learning capability of language model, integration of high quality, fine granular feedback from medical experts results in significant improvement in the performance of the language model.

The integration of expert feedback together with reinforcement learning method or reinforcement learning with human feedback (RHLF) has been successfully used to improve instruction following in LLMs [23]. Our approach to incorporate expert feedback in prompt tuning results in significant improvement in the quality of results however, the study does not evaluate the specific type of feedback from medical experts that is most effective in improving the accuracy of results. Specifically, medical expert input in the form of a structured, stepwise description may further improve the performance of the LLM as demonstrated in other domains using the Chain of Thought (CoT) approach. Further, the size of the study cohort in this paper is small; therefore, we are unable to draw broader conclusions regarding the effectiveness of the combined multiview and MEAG prompting framework. As part of our ongoing work, we aim to increase the size of the study cohort and analyze data from a broader spectrum of ophthalmology disorders using the new prompting framework introduced in this study.

## Data Availability

Available upon reasonable request.

## Acknowledgement

This research was funded in part by grants from the US National Institutes of Health (NIH): U24EB029005, R01DA053028, the US Department of Defense (DoD) grant W81XWH2110859, and the Clinical and Translational Science Collaborative of Cleveland, which is funded by the NIH, National Center for Advancing Translational Sciences, Clinical and Translational Science Award grant, UL1TR002548. The content is solely the responsibility of the authors and does not necessarily represent the official views of the NIH. Additional funding came from NEI T32: 5 T32 EY 24236-4, Case Western Reserve University Biomedical Research Fellowship-Hartwell Foundation (F.G.), Blind Children’s Foundation (F.G.), Research to Prevent Blindness Disney Amblyopia Award (F.G.), and Cleveland Clinic RPC Grant (F.G.). Support also included a Cleveland VA Medical Center grant 5I21RX003878-02 (AS and FG), Research to Prevent Blindness, CCLCM Unrestricted Block Grant, NIH-NEI P30 Core Grant, and Cleve-land Eye Bank.

## Notes

### Competing Interest Statement

The authors have declared no competing interest.

### Author Declarations

This study was reviewed and approved by the Cleveland Clinic Institutional Review Board (IRB). Written informed consent was obtained from each participant or parent/legal guardian for this study as mandated by the Declaration of Helsinki. The patient data with protected health information (PHI) elements was securely stored at Cleveland Clinic and only the deidentified data was subsequently shared with Case Western Reserve University for analysis by the transformer deep learning model.

### Summary of Updates

Amblyopia is a neurodevelopmental visual disorder that affects approximately 3-5% of children globally and it can lead to vision loss if it is not diagnosed and treated early. Traditional diagnostic methods, which rely on subjective assessments and expert interpretation of eye movement recordings presents challenges in resource-limited eye care centers. This study introduces a new approach that integrates the Gemini large language model (LLM) with eye-tracking data to develop a classification tool for diagnosis of patients with amblyopia. The study demonstrates: (1) LLMs can be suc-cessfully applied to the analysis of fixation eye movement data to diagnose patients with amblyopia; and (2) Input of medical subject matter expertise, introduced in this study in the form of medical expert augmented generation (MEAG), is an effective adaption of the generic retrieval augmented generation (RAG) approach for medical applications using LLMs. This study introduces a new multi-view prompting framework for ophthalmology applications that incorporates fine granularity feedback from pediatric ophthalmologist together with in-context learning to report an accuracy of 80% in diagnosing patients with amblyopia. In addition to the binary classification task, the classification tool is generalizable to specific subpopulations of amblyopic patients based on severity of amblyopia, type of amblyopia, and with or without nystagmus. The model reports an accuracy of: (1) 83% in classifying patients with moderate or severe amblyopia, (2) 81% in classifying patients with mild or treated amblyopia; and (3) 85% accuracy in classifying patients with nystagmus. To the best of our knowledge, this is the first study that defines a multi-view prompting framework with MEAG to analyze eye tracking data for the diagnosis of amblyopic patients.

## References

1. McKean-Cowdin, R., et al., Prevalence of amblyopia or strabismus in asian and non-Hispanic white preschool children: multi-ethnic pediatric eye disease study. Ophthalmology, 2013. 120(10): p. 2117–24.

2. Brown, T., et al., Language models are few-shot learners. Advances in neural information processing systems, 2020. 33: p. 1877–1901.

3. Chowdhery, A., et al., Palm: Scaling language modeling with pathways. Journal of Machine Learning Research, 2023. 24(240): p. 1–113.

4. Reid, M., et al., Gemini 1.5: Unlocking multimodal understanding across millions of tokens of context. arXiv preprint arXiv:2403.05530, 2024.

5. Touvron, H., et al., Llama 2: Open foundation and fine-tuned chat models. arXiv preprint arXiv:2307.09288, 2023.

6. Singhal, K., et al., Large language models encode clinical knowledge. Nature, 2023. 620(7972): p. 172–180.

7. Sahoo, S.S., Plasek J.M., Xu H., Uzuner O, Cohen C., Yetisgen, M., Liu, H., Stéphane, M., Wang, Y., Large Language Models for Biomedicine: Foundations, Opportunities, Challenges, and Best Practices. Journal of the American Medical Informatics Association (JAMIA), 2024.

8. Shah, N.H., Entwistle, D., Pfeffer, M.A., Creation and adoption of large language models in medicine. Jama, 2023. 330(9): p. 866–869.

9. Thirunavukarasu, A.J., Ting, D.S.J., Elangovan, K., Gutierrez, L., Tan, T.F., Ting, D.S.W., Large language models in medicine. Nature medicine, 2023. 29(8): p. 1930–1940.

10. Saab, K., et al., Capabilities of Gemini Models in Medicine. arXiv preprint arXiv:2404.18416, 2024.

11. Team, G., et al., Gemini: a family of highly capable multimodal models. arXiv preprint arXiv:2312.11805, 2023.

12. Guu, K., et al. Retrieval augmented language model pre-training. in International conference on machine learning. 2020. PMLR.

13. Ghasia, F. and J. Wang, Amblyopia and fixation eye movements. Journal of the Neurological Sciences, 2022: p. 120373.

14. Subramanian, V., R.M. Jost, and E.E. Birch, A quantitative study of fixation stability in amblyopia. Invest Ophthalmol Vis Sci, 2013. 54(3): p. 1998–2003.

15. Shi, X.F., et al., Fixational saccadic eye movements are altered in anisometropic amblyopia. Restor Neurol Neurosci, 2012. 30(6): p. 445–62.

16. Niechwiej-Szwedo, E., L. Colpa, and A.M. Wong, Visuomotor behaviour in amblyopia: deficits and compensatory adaptations. Neural Plasticity, 2019. 2019.

17. Vadhera, R. and M. Sharma. Review of Amblyopia and Artificial Intelligence Techniques Used for Its Detection. in Proceedings of the Second International Conference on Information Management and Machine Intelligence: ICIMMI 2020. 2021. Springer.

18. Ting, D.S.W., et al., Development and validation of a deep learning system for diabetic retinopathy and related eye diseases using retinal images from multiethnic populations with diabetes. Jama, 2017. 318(22): p. 2211–2223.

19. Holmes, J.M. and M.P. Clarke, Amblyopia. Lancet, 2006. 367(9519): p. 1343–51.

20. Webber, A.L. and J. Wood, Amblyopia: prevalence, natural history, functional effects and treatment. Clin Exp Optom, 2005. 88(6): p. 365–75.

21. Wang, Y., et al., Generalizing from a few examples: A survey on few-shot learning. ACM computing surveys (csur), 2020. 53(3): p. 1–34.

22. Research, S. EyeLink 1000 Plus: A highly accurate, precise, and versatile eye tracker. 2024; Available from: https://www.sr-research.com/eyelink-1000-plus/.

23. Ouyang, L., et al., Training language models to follow instructions with human feedback. Advances in neural information processing systems, 2022. 35: p. 27730–27744.

